# Non-contiguous Computed Tomography Lung Scans Can be Manipulated to Permit Artificial Intelligence Analyses for Interstitial Lung Disease in Systemic Sclerosis

**DOI:** 10.1101/2025.10.14.25338002

**Authors:** Murray Baron, Quan Nguyen, Bojan Kovacina, Charmaine van Eeden, Georg Langs

## Abstract

**Background:** Artificial Intelligence can analyse high resolution CT lung scans (HRCT) in various interstitial lung diseases (ILD) including Systemic Sclerosis (SSc). Older HRCT lung scans may have been saved as small dicom file sets consisting of non-contiguous slices. These are not amenable to AI analyses.

**Objectives:** Our aim was to develop and test a method of rebuilding small non-contiguous sets of HRCT lung slices into larger sets of contiguous slices that could be analysed by AI programs.

**Methods:** We deleted sets of dicom files from 14 large dicom file set scans from SSc patients and were left with a scan with about 30 equidistant non-contiguous slices. We then inserted copies of scans between each pair of slices to create a large dicom file set similar in size to the original large file set scan. Both the original scan and the rebuilt large dicom file set scan were analyzed by Contextflow ADVANCE Chest CT. We recorded the values for honeycombing (HC), reticular pattern (RP), ground glass opacities (GGO) and total ILD. We analyzed agreement between the original scan and the rebuilt large file set scan using intraclass correlation coefficient (ICC), Lin’s concordance correlation coefficient (CCC),^1^ Bland-Altman limits-of-agreement (LOA) plots and the Bradley-Blackwood p value.

**Results:** ICC, CCC, Bradly-Blackwood p values and Bland Altman plots showed excellent agreement between scans for HC, RC, GGO and total ILD except for the Bradley-Blackwood p value for RP.

**Conclusions:** Small non-contiguous HRCT lung scans in SSc can be manipulated to allow analysis by AI.

---

Interstitial lung disease (ILD) is a serious complication in systemic sclerosis (SSc, scleroderma) as a cause of significant morbidity and mortality.^2,3^ Computed tomography (CT) of the lung is the standard for diagnosing SSc-ILD and is also one component of following the trajectory of the disease.^4–9^ The extent of ILD at one timepoint has also been used in studies of associations with future deterioration of SSc-ILD.^10–16^

Machine learning CT quantitative evaluation of various aspects of ILD has become an important tool in the assessment of ILD and has been used for SSc-ILD. ^4,6,17–26^ Most such artificial intelligence (AI) algorithms have been designed to assess high resolution CT scans (HRCTs) with slice thickness < 2 mm and either contiguous or overlapping slices throughout the lung field.^26–28^ Recent HRCT lung studies fulfill these criteria with usually over 200 slices and one dicom file for each slice. We will refer to these as *large dicom files set scans*. Unfortunately, older HRCT lung studies included far fewer slices which were not contiguous and therefore could not be assessed by most AI programs. Most of these scans have only about 30 associated dicom files and we will refer to these as *small dicom file set scans*.

Because of our interest in SSc-ILD, we have collected HRCT scans from patients in the Canadian Scleroderma Research Group cohort, many of which go back to the early 2000s. Our goal is to have all these scans read by AI for further studies, but many of the older scans are composed of non-contiguous cuts with a total of only about 30 DICOM files per scan. As these cannot be read by most current AI algorithms, the specific objective of this study was to develop and test a method that would allow us to manipulate *small dicom file set scans* from HRCT lung scans in patients with SSc-ILD such that they could be successfully assessed by AI algorithms. Our hypothesis was that filling in the gaps of *small dicom file set scans* with copies of the scans on either sides of the gap would yield a l*arge dicom files set scan* that could be analysed by an AI algorithm and that the results would be accurate.

## Methods

### Overview of Study

We first developed an algorithm to build up non-contiguous *small dicom file* set scans into contiguous large dicom *files set scans*. The algorithm was created in Python and was designed to be run in Jupyter such that any user could convert *small dicom file set scans* into *large dicom files set scans*. We then obtained original *large dicom files set scans* from SSc patients of author MB with variable degrees of SSc-ILD. We deleted sets of DICOM files between slices from the CT scan that we wished to keep such that the resulting *small dicom file set scans* had only about 30 slices, mimicking older original scans with small dicom file sets (Figure 1). After the deletion of the sets of DICOM files, the remaining distance between each pair of slices was the same for all pairs as it is in older *small dicom file set scans*. We then used our algorithm to build those *small dicom file set scans* into what we will call *rebuilt large dicom files set scans* and both those and the original large dicom files set scans were analyzed by AI and the results of these pairs of scans were assessed for agreement with each other.

**Figure 1:**
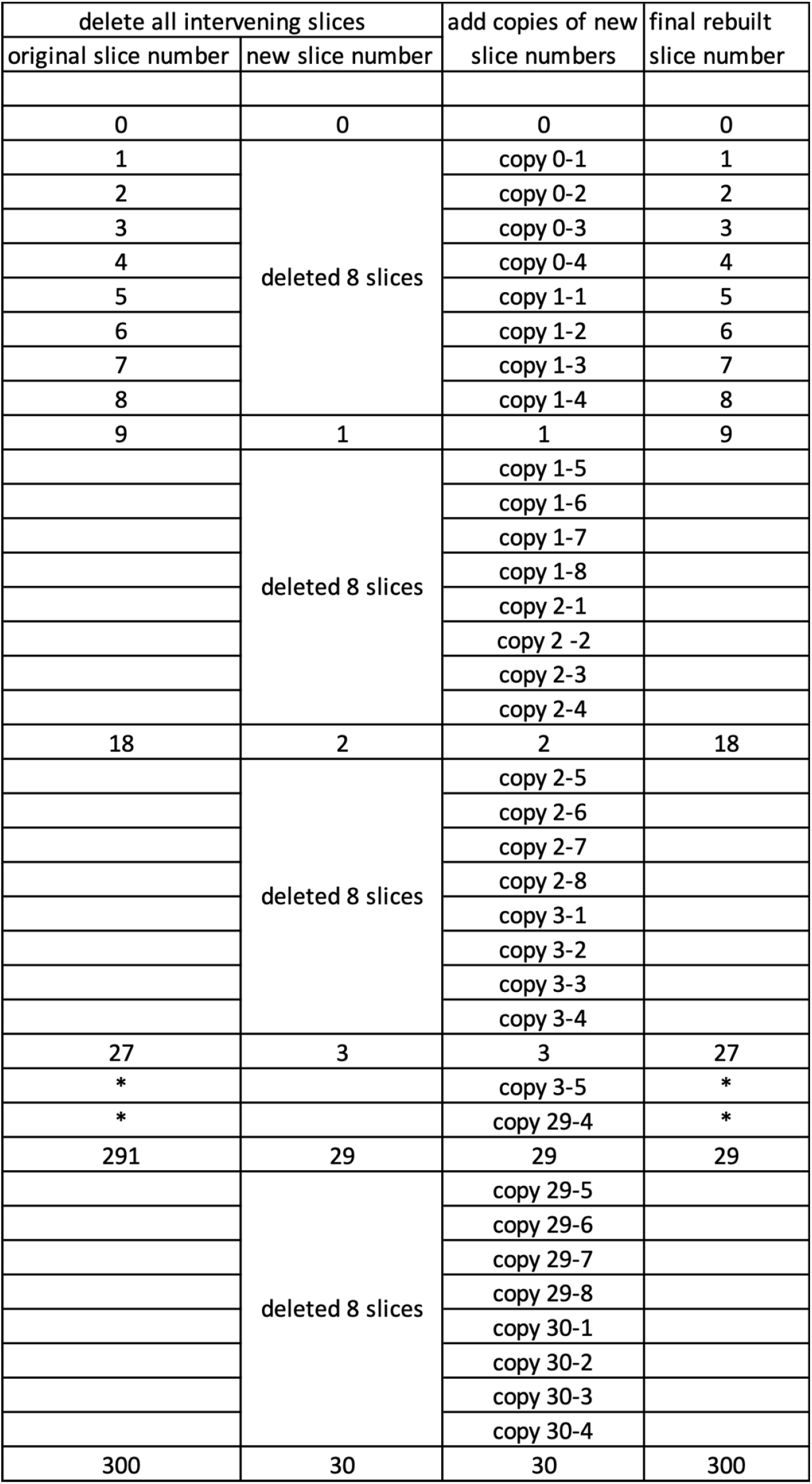
Illustration of how a large dicom file set CT scan was reduced to a small dicom file set and then rebuilt to final large dicom file set.

### Creating *small dicom file set scans* from original *large dicom files set scans*

We chose 14 *large dicom files set scans* from scans of our patients at the Jewish General Hospital, McGill University, Montreal. To ensure that our final set of scans covered various degrees of ILD, a chest radiologist (author BK) read all candidate scans and assessed the global percent of lung involved by ILD. The categories (and numbers of eventual chosen scans) were no ILD (3 scans), <10% (3 scans), 10-20% (2 scans), 20-30% (3 scans), or > 30% (3 scans).

Author MB reviewed each scan and deleted dicom files such that the resulting scan had about 30 non-contiguous slices equidistance apart (Figure 1).

### Algorithm For Rebuilding small dicom file set scans

We hypothesized that inserting copies of scans between each pair of slices would approximate what an original *large dicom files set scan* would look like. Depending on the slice thickness and the distance between the small files set slices, we copied each of the pair of slices and placed half of the total number of interval slices as copies of the first original slice and half as copies of the second original slice (Figure 1). So if we name the subsequent slices as 1, 2, 3 etc., and assuming that we needed X slices to be placed between each pair, we copied slice 1 and placed X/2 copies in order after slice 1 and we copied slice 2 and placed X/2 copies before slice 2. We repeated this for each set of pairs of the small dicom files set. In the end we now had a *rebuilt large dicom files set scan* with the same number of slices as the original *large dicom files set scan* and the slices were all contiguous.

The program to do this was written in python and automatically read the metadata from the dicom files of the *small dicom file set scan* and determined the slice thickness and the distance between the slices and calculated the number of copies to be made. It then changed the metadata of each copy to indicate its exact position in the order of slices to fulfill the requirements above.

The programmer made the program available in Jupyter with a set of instructions for the user explaining where the *small dicom file set scans* should be stored and how to adapt the code to allow the program to find those scans. After the script was run the rebuilt *large dicom files set scans* appeared in the same directory where the *small dicom file set scans* were stored.

### AI analysis of scans

Contextflow ADVANCE Chest CT is a deep learning-based, Computer Aided Detection (CADe) system for 3D medical imaging data. It uses convolutional neural networks (CNN) to detect and quantify pulmonary abnormalities, including emphysema, consolidation and reticulation by analysing volumetric chest CT data.^29,30^ The algorithm provides automated segmentation and classification of lung nodules and parenchymal changes, offering a quantitative insight into disease extent and severity. The AI model was trained using a diverse dataset of chest CT scans obtained from various patient populations covering a range of disease manifestations, including common and rare thoracic pathologies. To ensure high-quality training data, CT slices with and without pathologies were annotated and underwent multiple iterative quality control loops conducted by a team of expert radiologists. This annotation process involved precise labeling of pathological findings and abnormalities, ensuring the coverage of relevant features. The quality assurance steps included inter-observer validation, consensus reviews, and systematic refinements to ensure reliability of the training data. ^31^

### Data Analyses

The AI program assessed the overall percent of lung volume containing ground glass opacities (GGO), reticular pattern (RP), honeycombing (HC) or any ILD. We analyzed agreement between the AI analyses of the *rebuilt large dicom files set scan* and the original *large dicom files set scan* using Lin’s (1989) concordance correlation coefficient (CCC)^1^ to determine precision and accuracy, intraclass correlation coefficient (ICC) and Bland-Altman (1986) limits-of-agreement (LOA) plots to investigate agreement.^32^ These were computed using the Concord Module (Cox & Steichen)^33^ in Stata Statistical Software: Release 18, StataCorp (StataCorp, 2023). CCC determines the level of agreement between continuous measures obtained by two different methods or persons. The CCC increases when the data is tightly clustered near the reduced major axis (precision of the data) and when the reduced major axis is near to the line of perfect concordance (accuracy of the data), and this is graphically displayed as scatterplots (Cox & Steichen)^33^. The lower 95% CI were inspected, and values >0.8 were considered to show good agreement, and values >0.9 were considered to have excellent agreement. The Bradley-Blackwood p value compares the variance of the slope between the variables.^34^ p-values of <0.05 provide evidence of a lack of agreement between techniques. The agreement between the scales was assessed with the two-way mixed effects, absolute agreement, single rater intraclass correlation coefficient (ICC). Interpretations of ICC values for agreement differ but we have accepted < 0.5 as poor; 0.5 - 0.75 moderate; 0.75 - 0.9 good;> 0.90 excellent.^35^

## Results

The results of the of the AI analyses of the proportion of lung involved by HC, RP, GGO or total ILD of the original large file set and rebuilt large file set scans are in Table 1. The mean values for each of the lung abnormalities were very similar between the original and rebuilt scans (Table 2).

**Table 1.** AI analyses of rebuilt and original large DICOM file set scans.

| Case # | HC % | RP % | GGO % | ILD total |
| --- | --- | --- | --- | --- |
| 1 rebuilt | 0.00% | 1.63% | 4.52% | 6.15% |
| 1 original | 0.01% | 1.60% | 4.55% | 6.16% |
| 2 rebuilt | 0.17% | 2.16% | 6.75% | 9.08% |
| 2 original | 0.13% | 2.09% | 6.57% | 8.79% |
| 3 rebuilt | 0.28% | 1.02% | 9.22% | 10.51% |
| 3 original | 0.23% | 1.08% | 9.25% | 10.56% |
| 4 rebuilt | 0.02% | 0.15% | 0.39% | 0.56% |
| 4 original | 0.01% | 0.14% | 0.31% | 0.45% |
| 5 rebuilt | 0.05% | 1.42% | 21.72% | 23.19% |
| 5 original | 0.04% | 1.13% | 21.77% | 22.94% |
| 6 rebuilt | 9.30% | 3.83% | 4.97% | 18.11% |
| 6 original | 9.76% | 4.40% | 5.13% | 19.29% |
| 7 rebuilt | 0.01% | 1.09% | 66.09% | 67.18% |
| 7 original | 0.01% | 1.16% | 66.10% | 67.26% |
| 8 rebuilt | 5.15% | 3.85% | 4.98% | 13.98% |
| 8 original | 4.34% | 4.11% | 5.24% | 13.69% |
| 9 rebuilt | 0.02% | 0.25% | 0.52% | 0.78% |
| 9 original | 0.01% | 0.18% | 0.29% | 0.48% |
| 10 rebuilt | 12.43% | 3.03% | 2.35% | 17.80% |
| 10 original | 12.11% | 3.41% | 2.39% | 17.90% |
| 11 rebuilt | 0.00% | 0.09% | 0.52% | 0.61% |
| 11 original | 0.02% | 0.11% | 0.39% | 0.52% |
| 12 rebuilt | 0.03% | 0.23% | 0.29% | 0.54% |
| 12 original | 0.03% | 0.20% | 0.13% | 0.36% |
| 13 rebuilt | 0.47% | 1.76% | 0.61% | 2.85% |
| 13 original | 0.41% | 2.08% | 0.77% | 3.27% |
| 14 rebuilt | 0.01% | 0.26% | 3.37% | 3.64% |
| 14 original | 0.00% | 0.24% | 3.34% | 3.57% |

HC: honeycombing; RP: reticular pattern; GGO: ground glass opacities

**Table 2:** Comparison of ‘original’ and ‘rebuilt’ lung pattern analyses as percent of total lung volume.

|  | **‘original’**<br>**Mean (SD)** | **‘rebuilt’**<br>**Mean (SD)** | **Difference between**<br>**original and rebuilt**<br>**p-value** |
| --- | --- | --- | --- |
| Honeycombing | 1.936 (4.004) | 1.994 (4.039) | 0.969 |
| Reticular Pattern | 1.565 (1.486) | 1.482 (1.323) | 0.876 |
| Ground Glass | 9.015 (17.378) | 9.021 (17.361) | 0.999 |
| Overall ILD % | 0.125 (0.046) | 0.124 (0.046) | 0.997 |

Four scans had readings for total ILD of < 1 % which we interpret as no ILD. It can be seen visually that the values for all the pairs are very close. It can also be seen that there tended to be overestimation of total percent of lung involvement by ILD by the radiologist compared to the AI assessments as he rated 6 scans to have >20% overall ILD whereas the AI analyses detected only 2 scans in that category.

The intraclass correlation coefficient (ICC) values for all three lung measures (HC, RP, GGO), as well as overall ILD percentage, were excellent with all ICCs being >0.987 (Table 3). Concordance correlate coefficient (CCC) values for all four measures were also excellent, with all CCC values being >0.986. This suggests a high level of agreement between the analyses of the ‘original’ and ‘rebuilt’ scans. Although both ICC and CCC were excellent for RP, the Bradley-Blackwood p-value demonstrated a lack of agreement (p=0.004) for that particular abnormality (Table 3) due to larger differences in the mean and standard deviation for this measure (Table 3). High levels of agreement were confirmed for Honeycombing, Ground Glass and ILD % measures, with Bradly-Blackwood p values of >0.669 (Table 3).

**Table 3.** Concordance correlation coefficient analysis comparing ‘Original’ to “Rebuilt”.

| **Variables** | **ICC (95% CI)** | **CCC (95% CI)** | **Mean Difference**<br>**(95% LOA)** | **Bradley-**<br>**Blackwood**<br>**p-value\*** |
| --- | --- | --- | --- | --- |
| Honeycombing | 0.997 (0.993 -0.999) | 0.998 (0.995 – 1.000) | -0.058 (-0.583 – 0.467) | 0.669 |
| Reticular Pattern | 0.987 (0.961 -0.995) | 0.986 (0.974 - 0.997) | 0.084 (-0.352 - 0.519) | **0.004** |
| Ground Glass | 0.999 (0.999 – 0.999) | 1.000 (1.000 – 1.000) | -0.006 (-0.286 - 0.274) | 0.911 |
| Overall ILD % | 0.999 (0.999 – 0.999) | 1.000 (0.999 – 1.000) | -0.000 (-0.008 - 0.007) | 0.825 |

\*Values for p >0.05 indicate agreement

The scatterplots comparing ‘original’ and ‘rebuilt’ for measures HC, GGO, and total ILD indicated excellent precision and accuracy between the two methods, with near identical slopes and little spread of data away from the axis (Figure 2B, 2F, 2H). The scatterplot for RP indicated a scale shift, as indicated by the spread of data away from the reduced major axis and difference in the slopes of the reduced major axis and line of perfect concordance, suggesting only moderate accuracy. The scale shift resulted in values >1 being measured as smaller by the ‘rebuilt’ method (Figure 2D).

**Figure 2.**
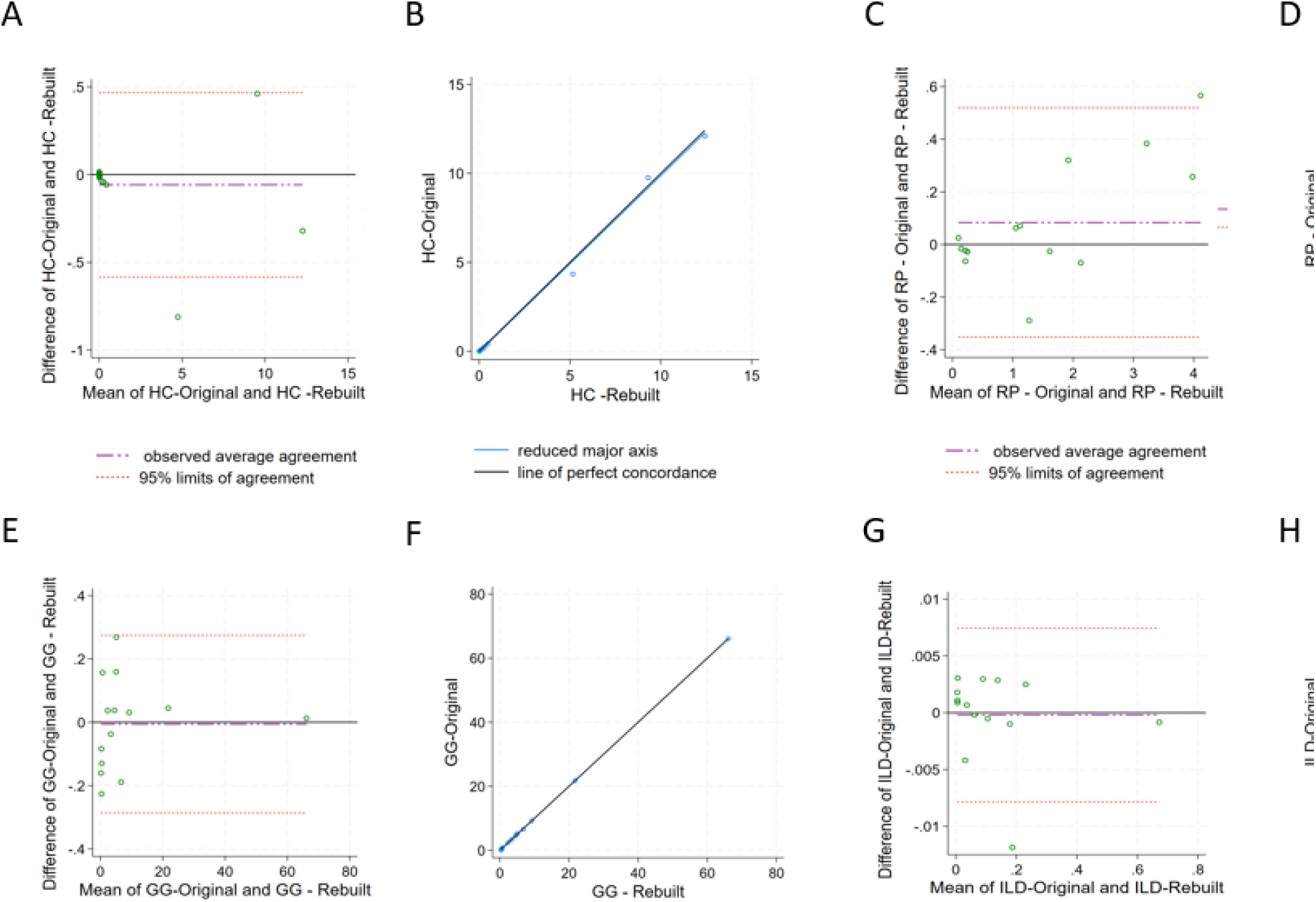
Graphs of agreement and concordance for CT measures using the Original and Rebuilt methods. A, C, E, G: Bland-Altman plots. The x-axis of the plot displays the mean measurement of the two instruments ((original and rebuilt scans) and the y-axis displays the difference in measurements between the two instruments. Agreement was assessed for the proportion of total lung volume occupied by honeycombing (HC), reticular pattern (RP), ground glass opacity (GG), or total Interstitial Lung Disease (ILD). The mean difference is presented as the long dashed horizontal line, no difference (mean difference = 0) is presented as the solid gray line and the limits of agreements (LOA) (± 1.96 SD) as the dashed lines. B, D, F, H: Scatter plots representing the line of perfect concordance (y = x) (dark gray) and the reduced major axis (light gray) corresponding to the regression line between the two measurements.

Agreement between the ‘original’ and ‘rebuilt’ methods is also visualized in Bland-Altman plots (Figure 2 A, C, E, G). Limits of agreement (LOA) (mean difference ± 1.96 SD) approximated 95% confidence. The mean difference for ILD (−0.000) and GGO (−0.006) was low, with narrow LOAs indicating good agreement (Figure 2E, 2H). The LOA for HC were wider, though constant around the mean, with a mean difference of −0.058 (Figure 1A). RP also had wider LOAs, with a mean difference of 0.084 (Figure 2C). For RP values data was more spread out from the mean, with differences between the two methods increasing as RP increased. ‘Rebuilt’ appears to underestimate RP values in the upper end of the mean (Figure 2C). This finding is consistent with the findings from the CCC analysis.

## Discussion

The particular objective of this exercise was to determine if we could have older HRCT scans from our cohort of SSc analysed in the future by AI methods. We have many scans going back as far as the year 2000 and the older scans are represented by small sets of DICOM files because the cuts are non-contiguous even though they are high resolution. These are not analyzable by most AI algorithms which are developed on large DICOM file sets from contiguous slices scans. We hypothesized that we could rebuild small DICOM file set scans into larger file set scans, where all slices were contiguous but the gaps were filled with copies of the original slices, and that the AI analyses would be accurate.

As it was not practical or ethical to request original scans from the same date on the same patient with both small and large DICOM file sets, we decided to start with large previously obtained DICOM file set scans, break those down to small DICOM file sets and then rebuild those to large DICOM file sets by filling in the gaps with copies (Figure 1). The metadata of each copy was changed to indicate its position in the new set of files.

Our results showed excellent agreement between the 2 scans for proportion of lung involvement by HC, RP, GG and total ILD. We performed agreement analyses by ICC, CCC, Bradley-Blackwood p values, Bland Altman plots and lines of concordance on scatter plots. All showed excellent agreement except for the analysis of RP especially by Bradley-Blackwood p value. As the proportions of RP involvement was always < 4.5%, we feel that any discordance between the values represented very small differences in absolute values and would thus not likely represent a significant clinical issue.

Assessing the proportion of lung involvement by total ILD alone is difficult to do visually. It can be seen that although our radiologist assessed 6 of the scans as having a total ILD of over 20% of lung volume, only 2 scans were assessed by AI as being in that category (Table 1). Assessing the proportion involvement by the sub-patterns of HC, RP and GG is even more difficult for a radiologist. AI has made these assessments much more reliable which will allow us to look for smaller changes over time more accurately and to better assess the contribution of the individual patterns to how patients feel and function and to assess the effects of various treatments on the different patterns.

For this study we have not yet analyzed the assessment of other lung changes that occur in ILD such as emphysema, bronchiectasis, vascular changes, airway wall thickness or others. Also, the importance of this work likely only applies retrospectively to researchers with large cohorts with older CT scans that wish to assess variables from those scans and their relationship to variables that were collected in those cohorts.

In summary, analyses by AI of ILD extent in older non-contiguous HRCT lung scans in SSc, and likely in other cases of ILD, is possible if the gaps between lung slices if filled in with copies of the adjoining original slices and the metadata of the new set of DICOM files is changed to reflect the new slice positions.

## Data Availability

All data produced in the present study are available upon reasonable request to the authors

## References

1. Lin LI. A concordance correlation coefficient to evaluate reproducibility. Biometrics 1989;45:255–68.

2. Wells AU, Steen V, Valentini G. Pulmonary complications: one of the most challenging complications of systemic sclerosis. Rheumatology (Oxford) 2009;48 Suppl 3:iii40–4.

3. Highland KB, Silver RM. Clinical aspects of lung involvement: lessons from idiopathic pulmonary fibrosis and the scleroderma lung study. Curr Rheumatol Rep 2005;7:135–41.

4. Goldin J, Elashoff R, Kim HJ, et al. Treatment of scleroderma-interstitial lung disease with cyclophosphamide is associated with less progressive fibrosis on serial thoracic high-resolution CT scan than placebo: findings from the scleroderma lung study. Chest 2009;136:1333–40.

5. Wells AU. High-resolution computed tomography and scleroderma lung disease. Rheumatology (Oxford) 2008;47 Suppl 5:v59–61.

6. Goldin JG, Lynch DA, Strollo DC, et al. High-resolution CT scan findings in patients with symptomatic scleroderma-related interstitial lung disease. Chest 2008;134:358–67.

7. Antoniou KM, Wells AU. Scleroderma lung disease: evolving understanding in light of newer studies. Curr Opin Rheumatol 2008;20:686–91.

8. Shah RM, Jimenez S, Wechsler R. Significance of ground-glass opacity on HRCT in longterm follow-up of patients with systemic sclerosis. J Thorac Imaging 2007;22:120–4.

9. Desai SR, Veeraraghavan S, Hansell DM, et al. CT features of lung disease in patients with systemic sclerosis: comparison with idiopathic pulmonary fibrosis and nonspecific interstitial pneumonia. Radiology 2004;232:560–7.

10. Denton CP, Goh NS, Humphries SM, et al. Extent of fibrosis and lung function decline in patients with systemic sclerosis and interstitial lung disease: data from the SENSCIS trial. Rheumatology (Oxford) 2023;62:1870–6.

11. Forestier A, Le Gouellec N, Béhal H, et al. Evolution of high-resolution CT-scan in systemic sclerosis-associated interstitial lung disease: Description and prognosis factors. Semin Arthritis Rheum 2020;50:1406–13.

12. Khanna D, Nagaraja V, Tseng CH, et al. Predictors of lung function decline in scleroderma-related interstitial lung disease based on high-resolution computed tomography: implications for cohort enrichment in systemic sclerosis-associated interstitial lung disease trials. Arthritis Res Ther 2015;17:372.

13. Le Gouellec N, Duhamel A, Perez T, et al. Predictors of lung function test severity and outcome in systemic sclerosis-associated interstitial lung disease. PLoS One 2017;12:e0181692.

14. Lopes AJ, Capone D, Mogami R, Menezes SL, Guimarães FS, Levy RA. Systemic sclerosis-associated interstitial pneumonia: evaluation of pulmonary function over a five-year period. J Bras Pneumol 2011;37:144–51.

15. Ramahi A, Lescoat A, Roofeh D, et al. Risk factors for lung function decline in systemic sclerosis-associated interstitial lung disease in a large single-centre cohort. Rheumatology (Oxford) 2023;62:2501–9.

16. Wu W, Jordan S, Becker MO, et al. Prediction of progression of interstitial lung disease in patients with systemic sclerosis: the SPAR model. Ann Rheum Dis 2018;77:1326–32.

17. Tashkin DP, Elashoff R, Clements PJ, et al. Cyclophosphamide versus placebo in scleroderma lung disease. N Engl J Med 2006;354:2655–66.

18. Tashkin DP, Elashoff R, Clements PJ, et al. Effects of 1-year treatment with cyclophosphamide on outcomes at 2 years in scleroderma lung disease. American journal of respiratory and critical care medicine 2007;176:1026–34.

19. Kim HG, Tashkin DP, Clements PJ, et al. A computer-aided diagnosis system for quantitative scoring of extent of lung fibrosis in scleroderma patients. Clin Exp Rheumatol 2010;28:S26–35. (Multicenter Study Randomized Controlled Trial).

20. Khanna D, Tseng CH, Farmani N, et al. Clinical course of lung physiology in patients with scleroderma and interstitial lung disease: Analysis of the scleroderma lung study placebo group. Arthritis Rheum 2011.

21. Kim HJ, Brown MS, Elashoff R, et al. Quantitative texture-based assessment of one-year changes in fibrotic reticular patterns on HRCT in scleroderma lung disease treated with oral cyclophosphamide. Eur Radiol 2011;21:2455–65.

22. Roth MD, Tseng CH, Clements PJ, et al. Predicting treatment outcomes and responder subsets in scleroderma-related interstitial lung disease. Arthritis Rheum 2011;63:2797–808.

23. Volkmann ER, Tashkin DP, Li N, et al. Mycophenolate Mofetil Versus Placebo for Systemic Sclerosis-Related Interstitial Lung Disease: An Analysis of Scleroderma Lung Studies I and II. Arthritis & rheumatology (Hoboken, NJ) 2017;69:1451–60.

24. Goldin JG, Kim GHJ, Tseng CH, et al. Longitudinal Changes in Quantitative Interstitial Lung Disease on Computed Tomography after Immunosuppression in the Scleroderma Lung Study II. Ann Am Thorac Soc 2018;15:1286–95.

25. Volkmann ER, Tashkin DP, Roth MD, Goldin J, Kim GHJ. Early Radiographic Progression of Scleroderma: Lung Disease Predicts Long-term Mortality. Chest 2022;161:1310–9.

26. Walsh SLF, De Backer J, Prosch H, et al. Towards the adoption of quantitative computed tomography in the management of interstitial lung disease. Eur Respir Rev 2024;33.

27. Lynch DA. Quantitative CT of fibrotic interstitial lung disease. Chest 2007;131:643–4.

28. Goldin JG. Quantitative CT of the lung. Radiol Clin North Am 2002;40:145–62.

29. Prayer F, Hofmanninger J, Weber M, et al. Variability of computed tomography radiomics features of fibrosing interstitial lung disease: A test-retest study. Methods 2021;188:98–104.

30. Pan J, Hofmanninger J, Nenning KH, et al. Unsupervised machine learning identifies predictive progression markers of IPF. Eur Radiol 2023;33:925–35.

31. Contextflow. Improving lung segmentation for higher coverage of clinically-relevant findings: a contextflow whitepaper [Internet. Accessed Available from: https://contextflow.com/2023/09/01/improving-lung-segmentation-for-higher-coverage-of-clinically-relevant-findings-a-contextflow-whitepaper/.

32. Giavarina D. Understanding Bland Altman analysis. Biochem Med (Zagreb) 2015;25:141–51.

33. Nicholas JC, Thomas S. CONCORD: Stata module for concordance correlation. S404501 ed: Boston College Department of Economics; 2000.

34. Russel C. Macro for Pairwise Reliability Indicators. The SouthEast SAS Users Group, Fourth Annual Conference. Atlanta, Georgia 1996:443–5.

35. Koo TK, Li MY. A Guideline of Selecting and Reporting Intraclass Correlation Coefficients for Reliability Research. J Chiropr Med 2016;15:155–63.

